# Evaluation of a Novel Intervention for Individuals at High Risk for Blindness from Diabetic Retinopathy: the SEEN Program Pilot

**DOI:** 10.64898/2026.10.29.26366801

**Authors:** Serina S. Applebaum, Akua Frimpong, Alvaro Granados, Julia J. Fu, Shannan Moore, Thomas Chang, Brittany Assanah-Lewis, Joana Andoh, June Weiss, Jingchen Liang, Yanhong Deng, Giselle Carlotta-McDonald, Ma’Glinda Ager, Lauren Kelley, Jacqueline Sanchez, Mayur Desai, Marcella Nunez-Smith, Josefa L. Martinez-Brockman, Kristen H. Nwanyanwu

## Abstract

**Objective:** To evaluate the feasibility and acceptability of the Sight-Saving Engagement and Evaluation in New Haven (SEEN) Program, designed to engage individuals at high risk for diabetic retinopathy (DR) in eye care.

**Study Setting and Design:** In this <u>single-arm, nonrandomized,</u> prospective intervention at a tertiary-care-affiliated institution in the Northeastern US, we enrolled 35 adults with diabetes at highest-quartile risk for DR, with no eye exam in the preceding year. Participants underwent comprehensive eye examination, DR education, and one year of social needs navigation by a community partner. We conducted a multi-method evaluation assessing program feasibility, acceptability, and preliminary clinical outcomes over 12 months. We thematically analyzed semi-structured interviews with participants, research team, and key partners. We organized themes using the RE-AIM, SEEN screening determinants, and Tailored Implementation in Chronic Diseases frameworks.

**Data Sources and Analytic Sample:** We collected surveys and electronic health record data from participants between July 2022 and September 2024. We conducted one-on-one interviews to understand the experience of the program between July 2024 and October 2024.

**Principal Findings:** We enrolled 26.5% of eligible individuals. Of 35 participants, 30 (85.7%) completed baseline social needs assessment, 25 (71.4%) completed the program, and 15 (42.9%) <u>had underwent</u> a follow-up eye exam. Patient navigators identified 31 social needs among 22 participants (62.9%). They spent a mean 96.8 (SD 43.8) minutes navigating participants over 12 months. Only two needs (6.5%) remained unresolved or unmet. HbA1c decreased among study completers (9.8 to 8.5; p=0.03). From 20 interviews, we identified 10 themes in 4 categories: feasibility (implementation, adaptive strategies, opportunities), acceptability (team dynamics, interpersonal interaction, procedural interface), appropriateness (external context, emotional context), and effectiveness (resources provided, lifestyle impact).

**Conclusions:** Pilot results support the feasibility and acceptability of the SEEN model to engage individuals at high risk for DR in eye care.

**Callout Box:** What is known on this topic

- Diabetic retinopathy is a leading cause of preventable blindness in the US, but screening adherence is suboptimal, particularly among marginalized populations.
- Patient navigation programs for other chronic diseases can facilitate engagement in care, and may be a component of successful programs to improve adherence to care for patients with increased support needs.

What this study found

- A program that risk-stratifies patients to provide high-touch education and social needs navigation shows promise for reducing barriers to diabetic retinopathy care and increasing eye exam utilization.
- Feedback from participants, key partners, and research team found the program feasible and acceptable

## Introduction

Diabetic retinopathy (DR) is a leading cause of preventable blindness in the US, and annual screening is recommended for individuals with diabetes.^1^ However, actual screening adherence is suboptimal, with disproportionately low rates among marginalized populations including Black, Latine, and low-income Americans.^2^ Despite evidence that early detection and treatment preserve vision, these groups often face delayed treatment and higher rates of vision loss.^3–5^ Addressing these disparities must be a public health priority to reduce preventable vision loss and improve health equity.

The disproportionate burden of blindness from DR is tied to social determinants of health, which affect healthcare access and outcomes.^6^ These factors create obstacles that prevent individuals from engagement and retention in eye care.^7^ Efforts to improve engagement must account for multifactorial barriers, ranging from unmet social needs to gaps in patient education. Patient navigation programs have shown promise in bridging these gaps for chronic diseases like diabetes and for cancer screening by providing tailored support to overcome logistical and structural challenges in healthcare access.^8,9^ However, there is limited evidence of the effectiveness of such programs in reducing disparities in DR screening specifically among high-risk populations.

The Sight-Saving Engagement and Evaluation in New Haven (SEEN) Program is a novel multi-level intervention designed to engage individuals at the highest risk for vision loss from DR in comprehensive eye care through risk stratification, clinical evaluation, education, and community-based care coordination. We present a patient-centered intervention designed to combat systemic barriers to health equity. We evaluate the feasibility and acceptability of the pilot SEEN Program to address the implementation gap between well-established screening guidelines for DR and the equitable access to DR screening in communities. Secondarily, we explore the effectiveness of the SEEN pilot.

## Methods

### Intervention

The SEEN Program pilot was a one-year program designed to engage individuals at high risk of blindness from DR in eye care.^10^ We identified individuals with diabetes through electronic health record (EHR) screen and community outreach. We applied a validated Cox model, the SEEN risk calculator,^11^ to identify those in the highest-quartile risk for DR progression and without an eye exam in over one year. Components of the SEEN risk calculator include having a primary care provider, health insurance, hemoglobin A1c (HbA1c), dyslipidemia, uncomplicated hypertension, and complicated hypertension (defined as history of hypertension and arrhythmia, transient ischemic attack, stroke, myocardial infarction, peripheral or pulmonary thrombosis/embolus, or heart failure). Enrolled participants underwent comprehensive ophthalmic examination, fundus photography, and point-of-care HbA1c testing. They viewed a co-designed educational video explaining the epidemiology, pathophysiology, and risks of DR.^12^ At the end of the eye exam, we directly connected participants with a local partner (Project Access-New Haven; PA-NH) for patient navigation and community health worker services. At that time, participants scheduled a follow-up phone call with PA-NH for in-depth baseline assessment (conducted a median of 7 days after initial contact). Trained PA-NH navigators screened participants for unmet social needs, and subsequently provided resources, check-ins, reminders, and coordination for health visits, focused on but not exclusive to DR screening and eye care.^13^ Patient navigators communicated primarily via telephone on a quarterly basis, or more frequently depending on individual participant needs <u>for one year</u>. Participants were compensated $20 at the start of the program, and $15 for each point of navigation up to a total of $100.

This is a single arm pilot study without a comparison group. In this study, we used a convergent multi-method design to integrate qualitative and quantitative data evaluating the process (feasibility, acceptability, and appropriateness) and outcomes (effectiveness) of the SEEN Program intervention.^14^ In the context of implementation, feasibility represents the degree to which we could implement the intervention among participants. Acceptability represents how satisfied participants were with the intervention. Appropriateness assesses the relevance of the intervention and its compatibility with participants’ needs. Effectiveness measures the intervention’s impact under real-world conditions.^15^

The SEEN Program is grounded in an integrated framework for determinants of DR screening developed by our team.^7,16^ The Tailored Implementation for Chronic Diseases (TICD) framework guided the strategic development of the program by identifying the determinants of implementation outcomes.^17^ The study protocol was approved by the Yale University Institutional Review Board. All participants provided written informed consent to participate and the study is in accordance with HIPAA regulations. The recruitment period began on July 25, 2022, and continued until September 30, 2023. This study is registered on ClinialTrials.gov, ID # NCT05188703, registered July 25, 2022. The protocol and statistical analysis plan can be accessed at the same URL. De-identified participant data are available upon written request to Dr. Kristen Nwanyanwu.

#### Part 1: Quantitative evaluation

We quantitatively assessed resource utilization and participant outcomes between July 2022 and September 2024.

### Data collection

We conducted surveys of participants’ sociodemographic and health characteristics. Variables not included in clinical assessment were abstracted from the EHR. We collected and managed study data using REDCap electronic data capture tools hosted at Yale University.^18,19^ We categorized participants as having completed an eye exam within 12 months after initial assessment (normal utilizers) or not returning for follow-up eye exam (under-utilizers). Baseline variables included age, sex, race and ethnicity, insurance coverage, and barriers to care (e.g., transportation, food security, housing). We collected systemic health measures including pre-existing and newly diagnosed comorbidities, laboratory values, and interval emergency department visits and hospitalizations. Ophthalmic health variables included presence and stage of DR, ocular comorbidities, visual acuity, intraocular pressure (IOP), and interval procedures or treatments. We reported IOP as the mean of each individual’s two eyes. Additionally, we recorded metrics of patient navigation success, including time spent per participant, number of navigator touchpoints, and number of unmet needs.

### Statistical analysis

We summarized continuous variables as means and standard deviation and categorical variables as counts and percentages, unless otherwise specified. We used the Wilcoxon signed-rank test and McNemar’s chi-square tests for continuous and categorical variables, respectively, to determine changes in systemic and ophthalmic outcomes from enrollment to 12 months. Participants completed the program if they attended the baseline eye exam and completed baseline social needs screening and all 4 quarterly PA-NH check-ins. We compared baseline characteristics between study completers and non-completers using independent samples t-tests and chi-square tests. We completed analyses using SAS version 9.4 (SAS Institute Inc, Cary, NC), with p<0.05 considered statistically significant.

#### Part 2: Qualitative evaluation

We conducted semi-structured interviews with study team members, key partners, and a purposive subset of program participants between July 2024 and October 2024.

### Interview guide design

We developed two complementary interview guides: one for program participants and a second for research team members and key partners. We leveraged our SEEN Framework for determinants of DR screening supported by the Consolidated Framework for Implementation Research (CFIR) and the Reach Effectiveness Adoption Implementation Maintenance (RE-AIM).^7,20,21^ We made minor revisions to both guides after three initial interviews each. Questions in the participant interview guide addressed experiences with the patient navigator, satisfaction with the program, and areas for improvement (Supplemental Table 1). The research team and key partner interview guide addressed similar components, in addition to perceptions of challenges patients face, the process of implementation, and program sustainability (Supplemental Table 2).

### Data collection

We interviewed a purposive sample of 10 research team members and key partner staff representing diverse roles (clinical technicians, patient navigation team, data management, program administration, research assistant, Principal Investigator) to ensure a comprehensive representation of perspectives relevant to the study. We also selected a subset of participants who completed the yearlong program. Interviews were conducted via telephone or video conferencing platform by members of the research team (A.F., S.A., A.G., S.M.). Interviews were audio recorded with informed consent and professionally transcribed. Interviews lasted 30 to 45 minutes. SEEN Program participants received $15 compensation for completing the interview. Research team members and key partners were not compensated for interviews.

### Interview analysis

We analyzed de-identified transcripts using NVivo software (v14, Lumivero, Denver, CO). Four authors independently analyzed each transcript: S.A. (White female, third year medical student), A.G. (Hispanic male, research associate), A.F. (Black female, fourth year medical student) and S.M. (Black female, fourth year medical student) with an approach informed by the RE-AIM framework and thematic analysis.^21,22^ We established preliminary codes for the participant interviews from a previously developed framework from the SEEN lab,^7^ which we expanded inductively during an initial pilot coding round of the first 3 interviews. The analysts independently coded each interview transcript, then discussed all discrepancies to reach consensus. After finalizing the codebook, we identified a set of emergent overarching themes. We conducted participant interviews until we achieved thematic saturation. We underwent a parallel process of coding and thematic analysis for the research team and key partner interviews to identify any discrepancy or additional themes to the participant interviews.

## Results

### Quantitative measures

#### Process outcomes (feasibility, acceptability, appropriateness)

A summary of the results is included in Table 1. From a composite total 1536 potential participants identified from all recruitment sources, 132 met full inclusion criteria to be eligible for participation. Of 132 eligible individuals, 64 (48.5%) were interested in participation after initial contact and 35 (26.5%) were ultimately enrolled (Figure 1).

**Figure 1.**
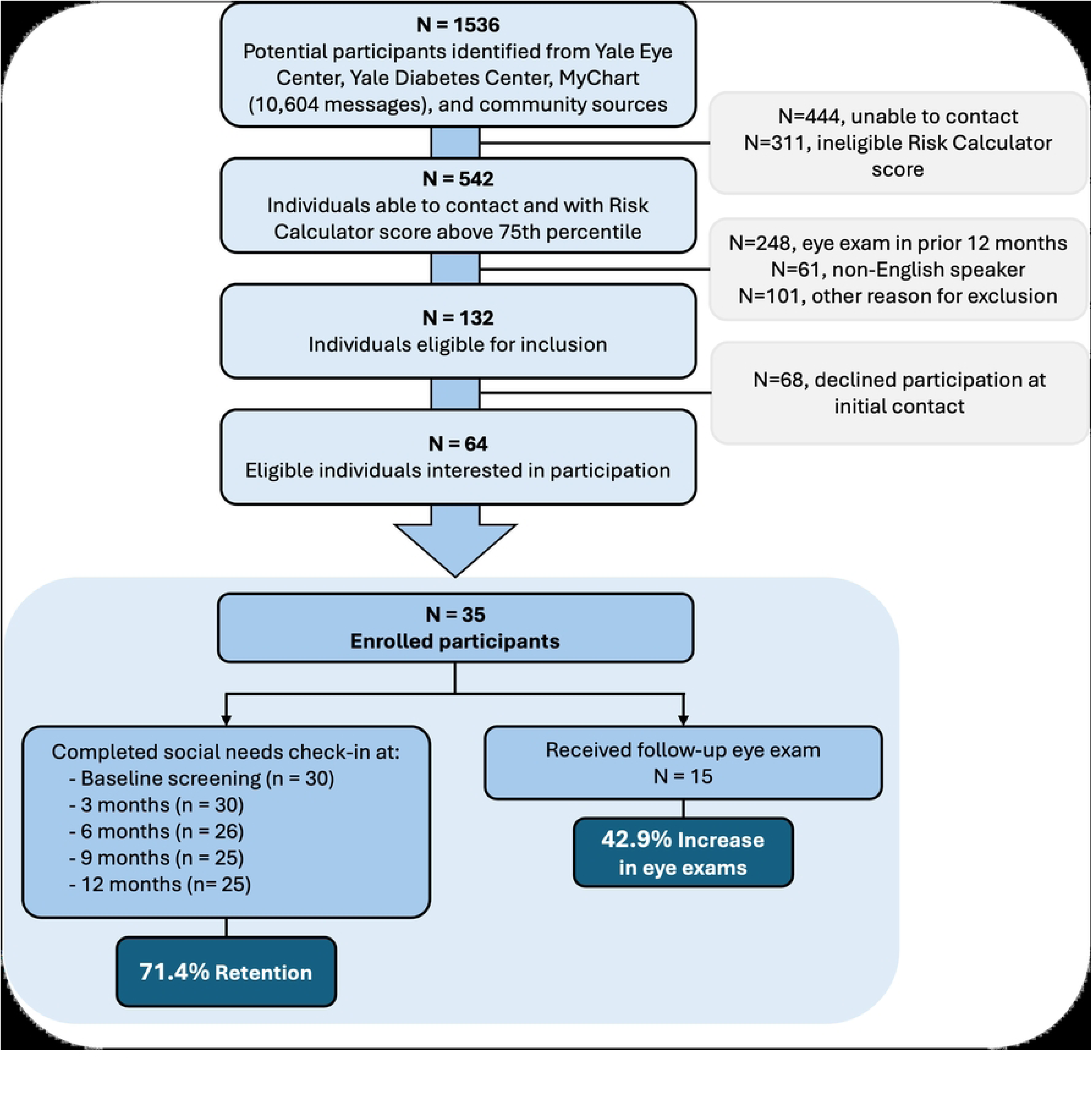
Participant selection flowchart.

**Table 1.**
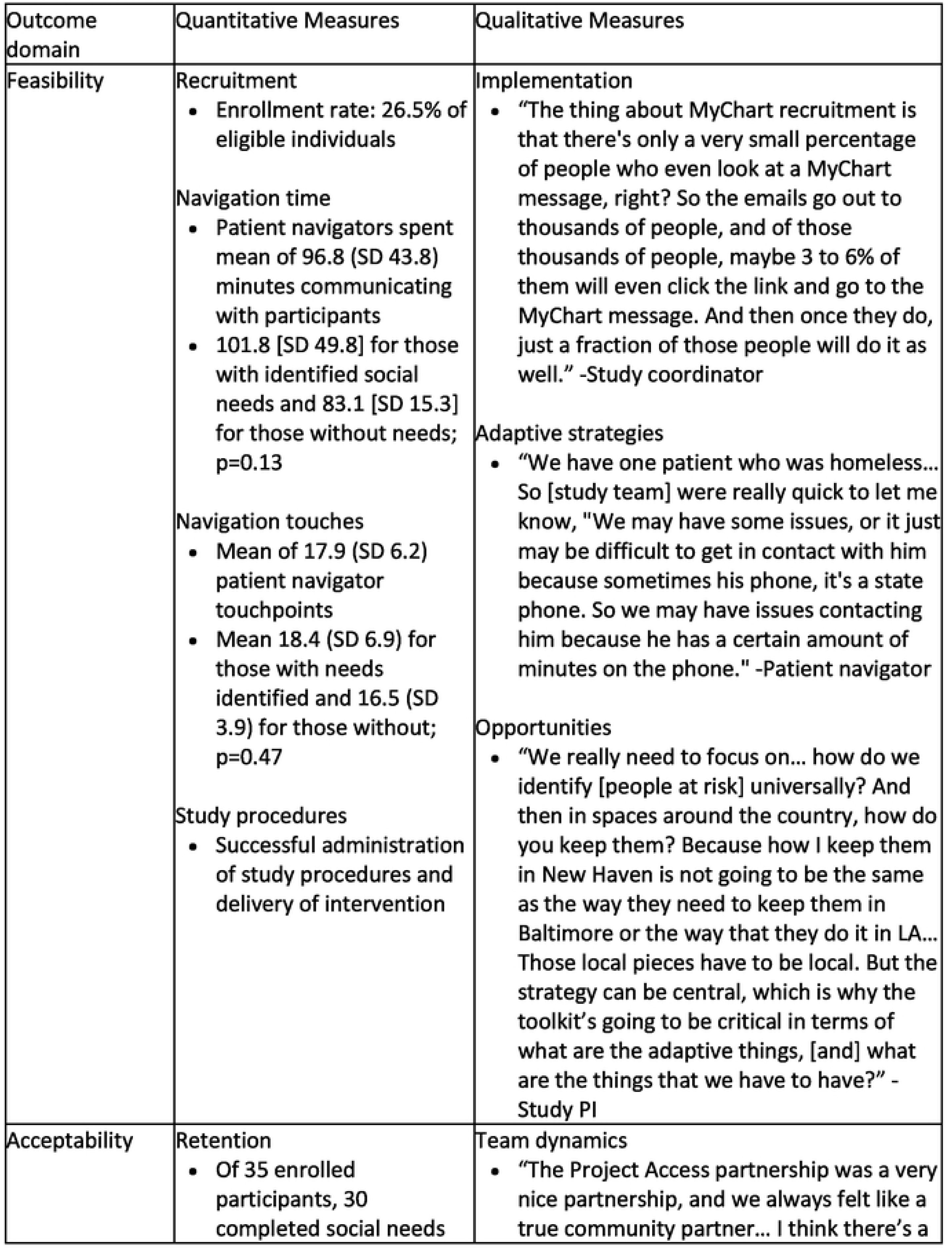
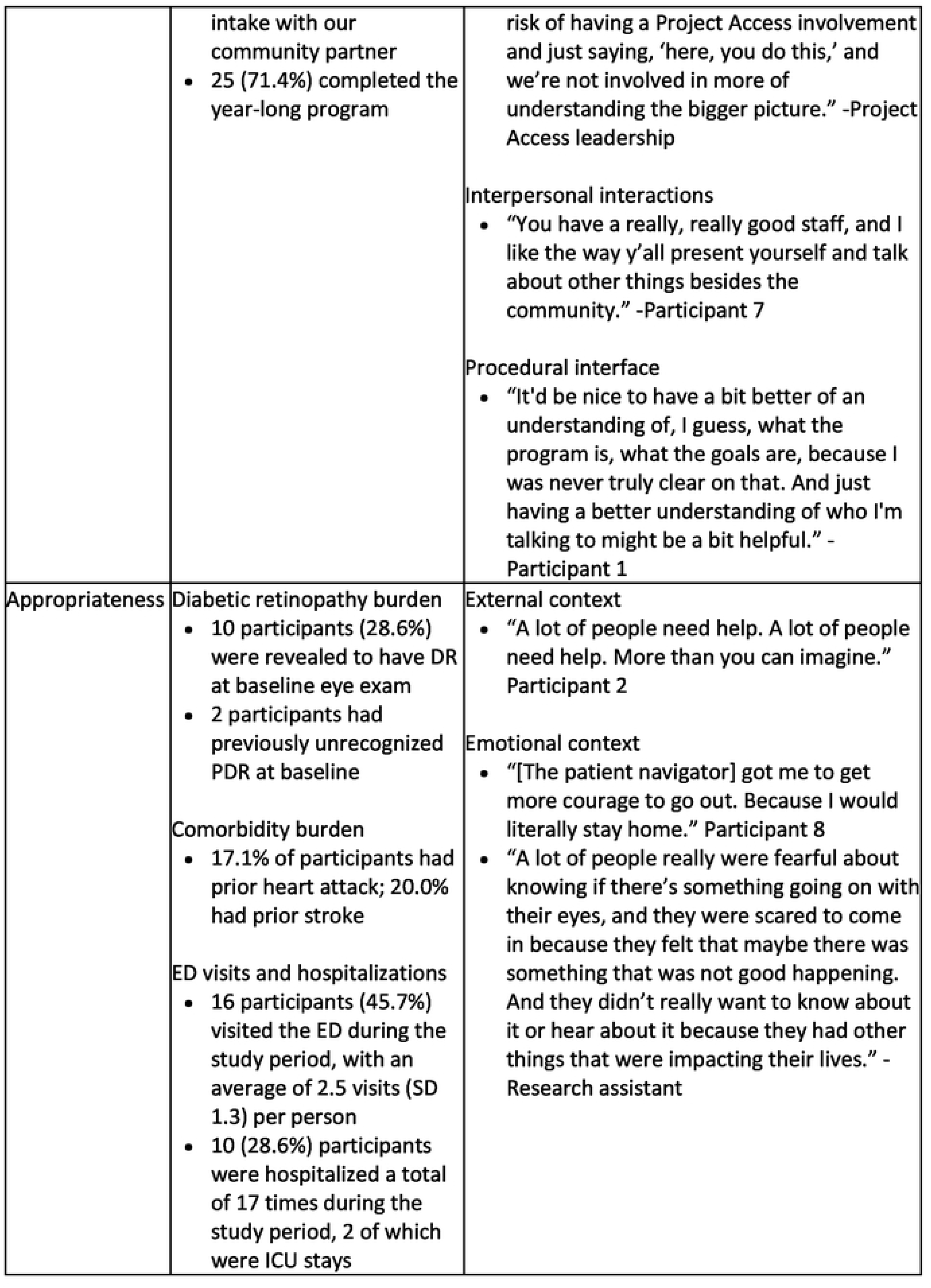
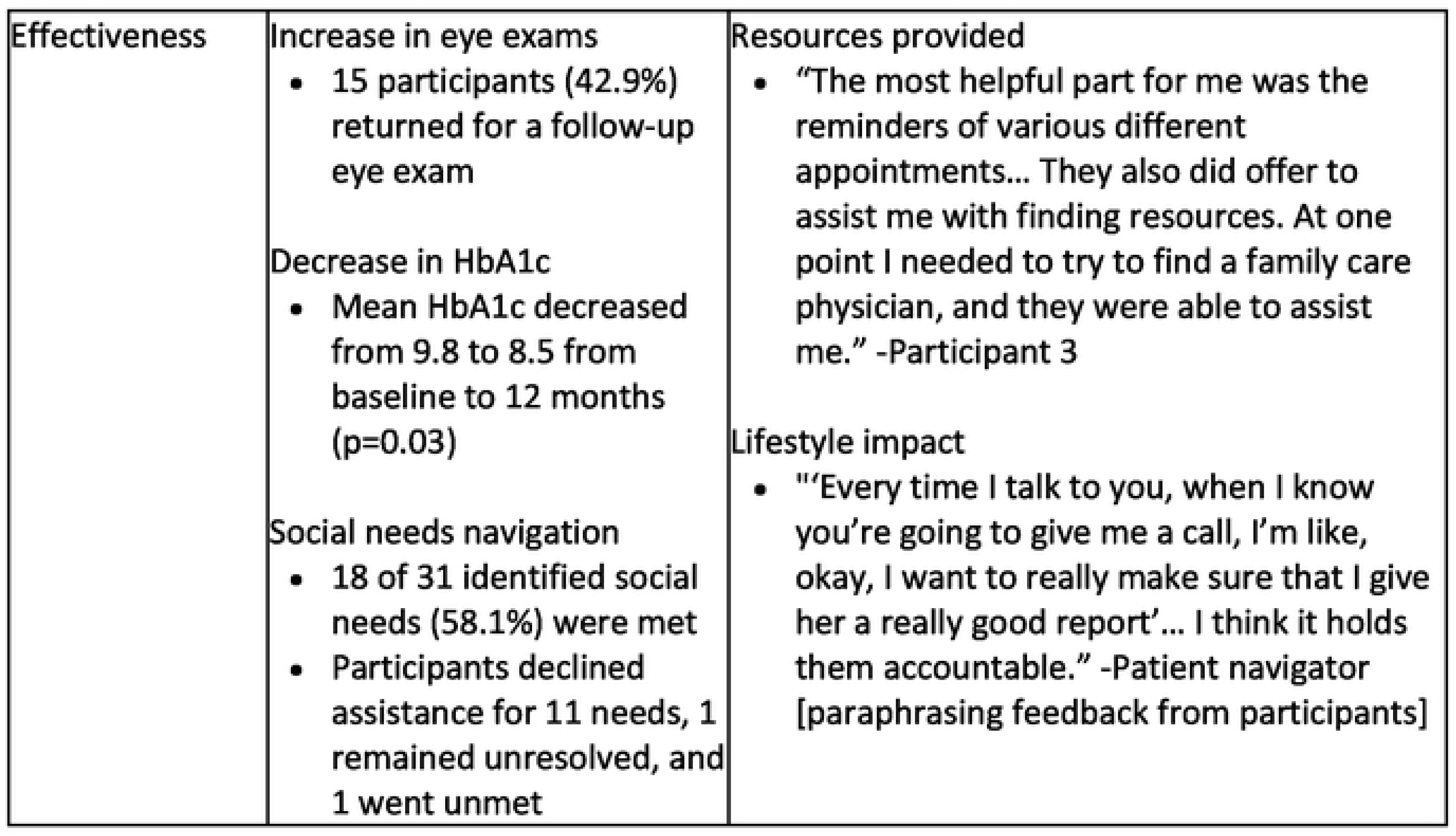
Summary of SEEN Pilot multi-method evaluation results in 4 different implementation outcome domains.

Of the 35 enrolled participants, 30 completed a social needs intake with our community partner and 25 (71.4%) ultimately completed the year-long program (Figure 1).

Individuals who completed the study trended toward higher baseline systolic and diastolic blood pressure than those who did not complete the study (p=0.06; p=0.09). There were no other differences between those who did and did not complete the study in baseline demographic, ophthalmic, or systemic health-related variables. Baseline characteristics and retention rates are shown in Supplemental Table 3.

Overall, patient navigators spent a mean of 96.8 (SD 43.8) minutes over 12 months communicating with participants (101.8 [SD 49.8] for those with identified social needs and 83.1 [SD 15.3] for those without identified needs; p=0.13). This included a mean of 18.4 touchpoints (SD 6.9) for those with needs identified and 16.5 (SD 3.9) for those without (p=0.47). The baseline screening was the most time-intensive timepoint, requiring a mean of 40.7 minutes (SD 21.0). Patient navigation required 35-39% full-time equivalent (FTE) effort from one navigator.

Regarding comorbidities, one participant developed end-stage renal disease, one developed a new diabetic foot ulcer, and two were newly diagnosed with congestive heart failure during the year following enrollment. Eighteen participants (51.4%) visited the emergency department, with an average of 2.6 visits (SD 1.5) per person. Chief complaints ranged from primary care needs like medication refills and cough, to emergencies such as hypoglycemia. Eleven participants (31.4%) were hospitalized a total of 17 times in the year since enrollment, 2 of which required intensive care.

#### Effectiveness outcomes

Among the 35 program participants, 15 (42.9%) were normal utilizers of eye care (at least one eye exam after the baseline exam). Among those who did and did not complete the study, 52% and 20%, respectively, had a follow-up eye exam (p=0.13) (Figure 2). The number of follow-up eye exams among normal utilizers ranged from 1-16 (median, 1). Most follow-up eye exams were with retina specialists or comprehensive ophthalmologists. Of the 20 participants who had no follow-up eye exam, 9 (45.0%) never scheduled follow-up at all, and 11 (55.0%) had at least one ophthalmology appointment scheduled but either canceled or did not attend (‘no-show’). Most (76% and 70%, respectively; p=0.69) participants who completed and did not complete the study had at least one cancelled or not attended visit (Figure 2).

**Figure 2.**
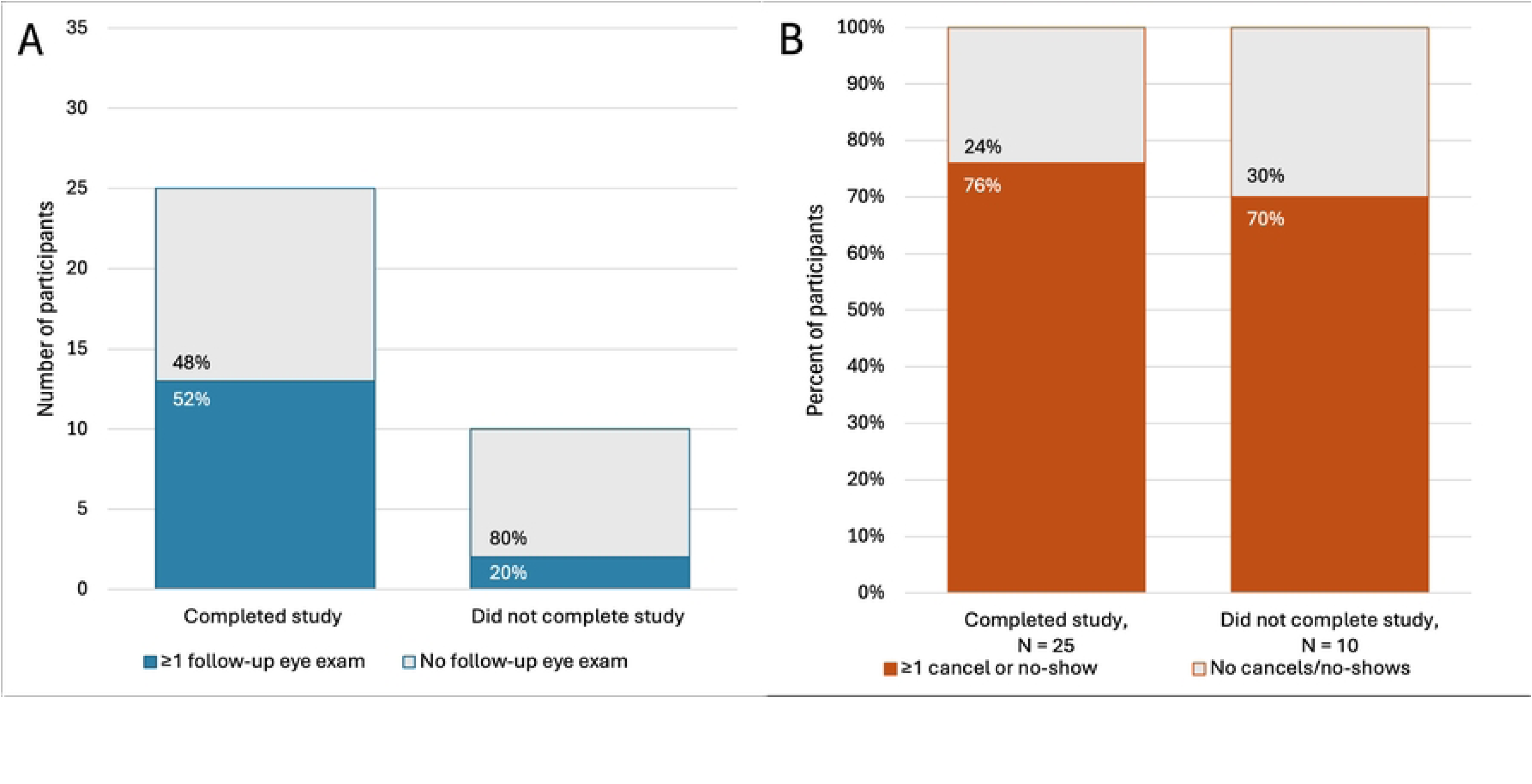
(A) number of participants with completed ophthalmology visits, and (B) proportion of participants with cancelled or no-show ophthalmology visits over 12 months since enrollment in the SEEN pilot study.

Among individuals who completed the study, mean HbA1c significantly decreased from baseline to 12 months (9.8 to 8.5; p=0.03), while visual acuity and intraocular pressure did not change (Table 2). In contrast, mean HbA1c from baseline to one year after enrollment was not significantly different for individuals who did not complete the study (p=0.36).

**Table 2.**
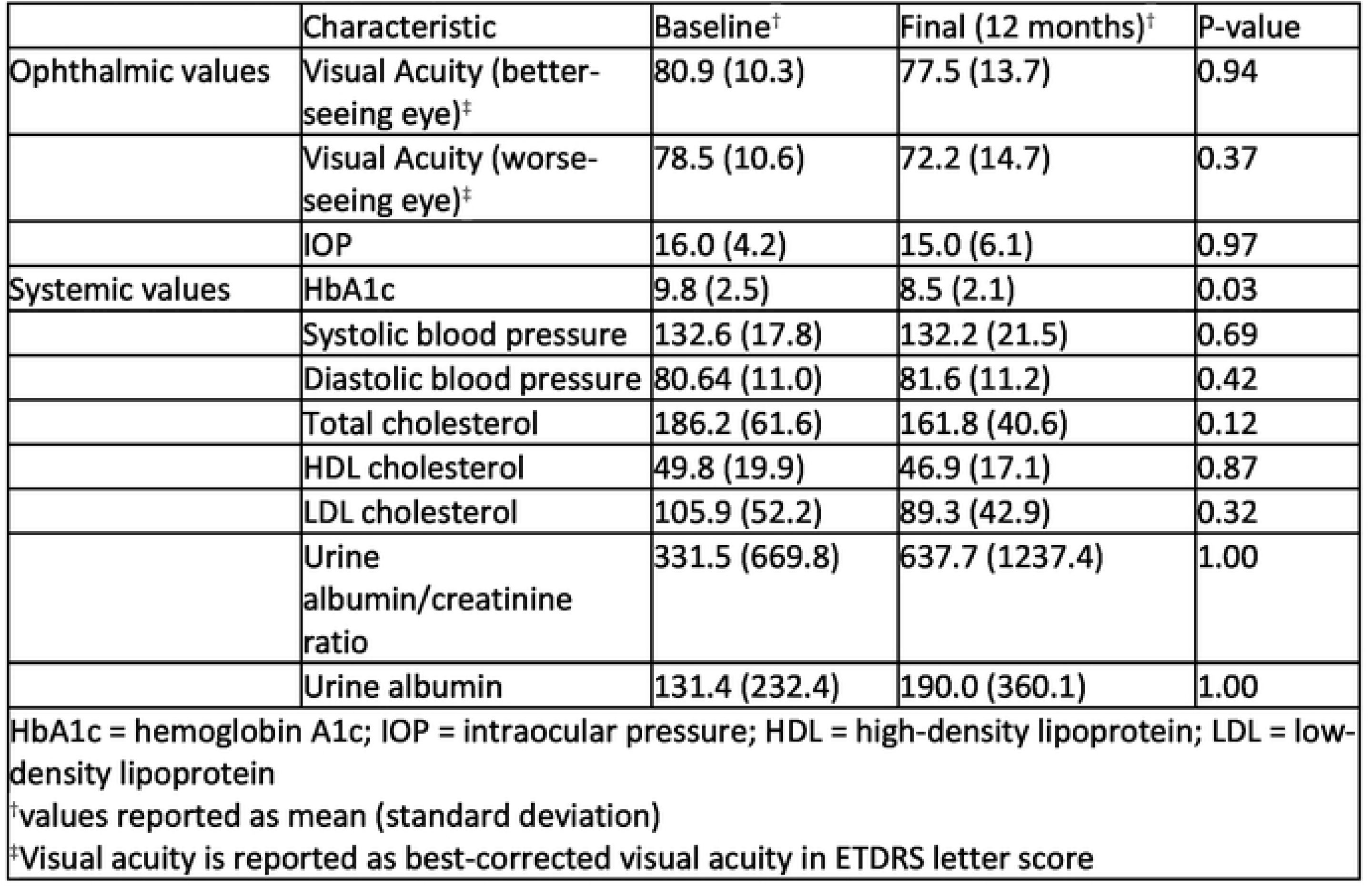
Change in clinical values over time among SEEN Program participants who completed 12 months of follow-up (N=25).

Three of 10 participants with DR on baseline exam returned for subsequent eye exams, at which all 3 individuals maintained stable DR stage. Two of these 3 (66.7%) had better visual acuity at the final eye exam. The remaining participant had high-risk proliferative DR at baseline and, during the study period, developed neovascular glaucoma and a tractional retinal detachment, ultimately requiring vitrectomy, multiple sessions of panretinal photocoagulation, and intravitreal injections of anti-vascular endothelial growth factor (anti-VEGF).

Our patient navigation community partner identified 22 participants (62.9%) with 31 total social needs in the categories of food access (n=19; 61.3%), transportation (n=6; 19.4%), primary care provider (n=4; 12.9%), health insurance (n=1; 3.2%), and housing (n=1; 3.2%). Eighteen of these 31 needs were ultimately met, while participants declined assistance for 11 needs, 1 need remained unresolved (i.e., need not actively addressed due to waitlist placement), and 1 participant’s need went unmet (i.e., need ongoing and could not be met). The instances of health insurance and housing needs were both met (Figure 3).

**Figure 3.**
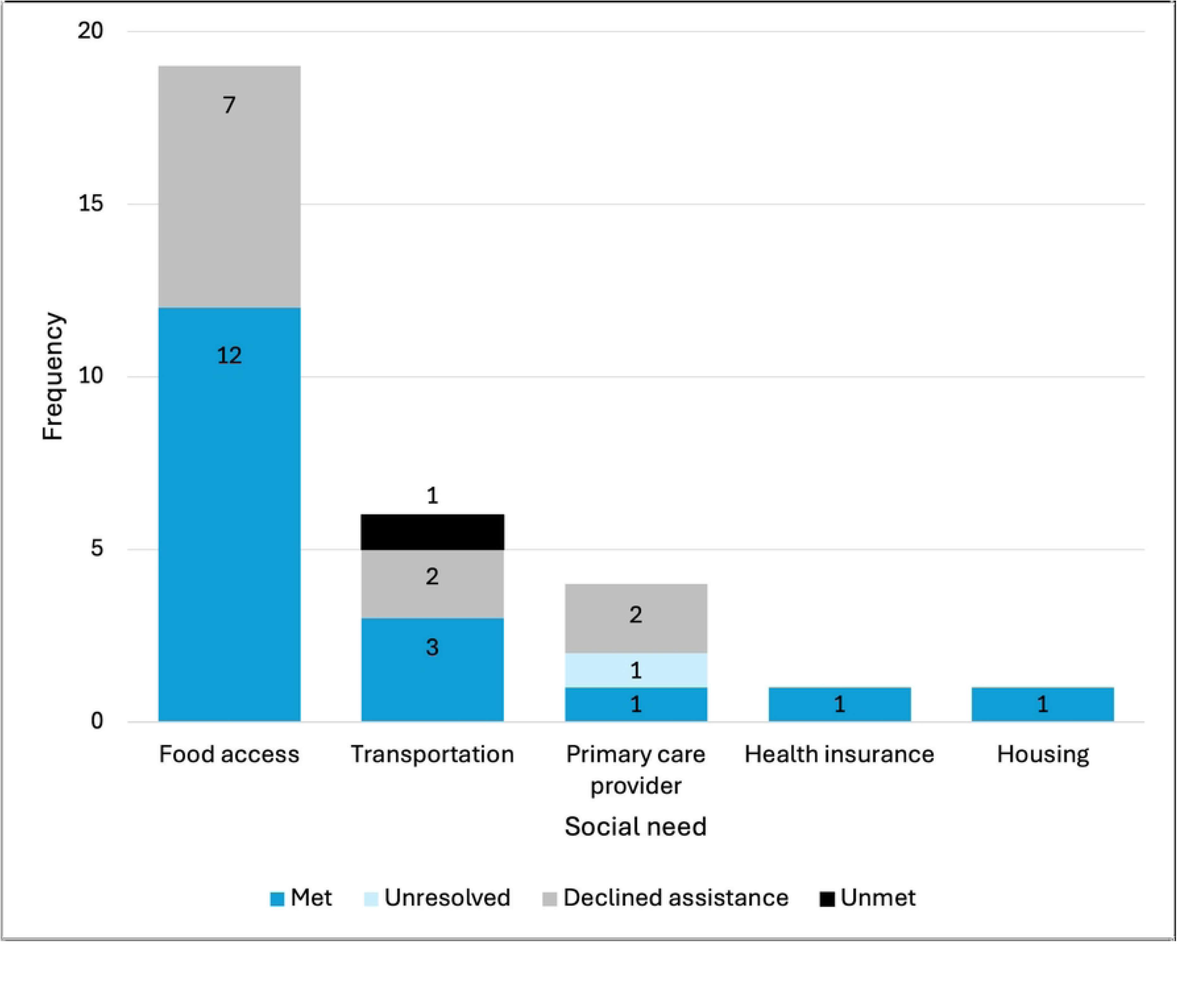
Outcomes of social needs identified by 30 participants in the SEEN Program pilot at 12 months after enrollment.

### Semi-structured interviews

We interviewed 10 of the 35 participants enrolled in the SEEN Program Pilot study and 10 members of the research team or key partner staff. The recording of one interview was not properly saved due to an inadvertent technical issue, necessitating a re-interview with that participant.

We identified 10 prominent themes, grouped into the 4 overarching categories of program feasibility, acceptability, appropriateness, and effectiveness (Table 1). Feasibility themes included overall implementation, adaptive strategies, and opportunities. Acceptability factors included team dynamics, interpersonal interaction, and participant’s procedural interface. We divided appropriateness into external and emotional context. Effectiveness themes included resources provided and lifestyle impact.

#### Feasibility

The study team described successes and challenges of program implementation, particularly in recruitment, communication, follow-up, and logistics. Initial recruitment efforts were hindered by the COVID19 pandemic and a primarily EHR messaging-based recruitment strategy, which yielded a low response rate. Other challenges included difficulties contacting participants, coordinating communication between research team and key partners, securing clinic space for eye exams, and distributing participant compensation.

Adaptability emerged as a central theme in overcoming hurdles. The program team tailored solutions to participants’ individualized needs, which were different for those with complex comorbidities or for one unhoused participant. They also reported diligence and flexibility required to adapt to unexpected events such as malfunctioning machinery, staffing challenges, and coordination of participants. “I was on a team that was really persistent. The team was amazing, and they just kept going no matter what and would keep trying and would try a different way…because they believe in the mission” said the study PI.

Opportunities identified by participants, research team, and key partners included strategies for program expansion, additional resources that participants might benefit from such as cooking classes and increased staffing, and structural changes to improve SEEN Program protocol flow such as timing of social needs screening.

#### Acceptability

The research team and key partners reported that collaborative team dynamics including strong support and communication between different branches of the team were helpful in achieving program goals. They also noted the value of enlisting domain experts, for example citing the patient navigation expertise of our key community partner, PA-NH, who were able to best serve participants through their longstanding experience.

Participants described their interpersonal interactions with the research team and key partners as helpful and empathetic. One participant appreciated the accessible communication style, saying “they talk like a normal person, not a doctor” (Participant 8).

Participants reflected on their own interface with study procedures. Some echoed communication challenges and confusion about the study’s goals. Our financial processing system created unanticipated changes midway through the pilot program. This caused delays in compensation, which two participants described as an inconvenience. Ultimately, most participants (90%) reported that they would recommend the program to someone else.

#### Appropriateness

Participants’ external context reinforced the need for the intervention. Participants described external circumstances that justified a need for support, including unemployment and disability, isolation from family, competing health concerns, and poor technology literacy. Participants and key partners observed a community-wide need for this program because of the high prevalence and severity of disease, and insufficient diabetes-related knowledge in the local community. One participant noted the complexity of the healthcare system, which necessitates additional navigation support for many people (Participant 1).

Emotional context also played a role in program implementation. One participant mentioned feeling “belittled” by researchers in the past, which previously dissuaded him from participating in studies (Participant 2). A common insight was the fear associated with diabetes, including discovering complications or becoming dependent on insulin.

Participants reported this fear can drive avoidance of healthcare. Conversely, participation in the study revealed that “diabetes is a controllable thing, and you don’t have to be scared of it” (Patient Navigator).

#### Effectiveness

We connected program participants to tangible resources including medical care, diet and exercise support, financial assistance, and transportation. Participants and the study team reported the positive impact of these resources on patients’ ability to engage with the healthcare system and manage their health. Importantly, the study team observed the high quality of care provided, especially the initial eye exam: “the care provided was the same level of care that anyone should expect to receive if they walk through the doors at the eye center” (Clinical Trial Director).

Lifestyle impacts included habit changes and increased health awareness facilitated by patient navigation. Participants described that the accountability provided by regular check-ins helped shape their mindset and behavior. One participant described, “the times when I would try to schedule a new appointment or check on my medical supplies were usually after one of those [patient navigator] calls because it was at the forefront of my mind” (Participant 1).

## Discussion

Whereas the current system oversimplifies DR screening by assuming uniform risk for all patients, the SEEN Program offers an opportunity to identify and intervene for those at highest risk. This multi-method evaluation showed that our pilot program integrating clinical evaluation, education, and navigation services for individuals at high risk for DR was both feasible and acceptable in a diverse Northeastern US city. Our findings also suggest evidence of effectiveness in improving eye care utilization and health outcomes. We successfully engaged over 70% of participants through one year of patient navigation, increased eye exam utilization, and observed a significant reduction in HbA1c levels.

Our target population of individuals at high risk for DR without recent eye exams is inherently disengaged from the healthcare system and therefore challenging to re-engage. Recruitment required multiple iterations. Initial outreach via electronic health record messaging was ineffective. Many individuals in the target population faced barriers such as limited internet access, difficulty navigating digital platforms, or inconsistent access to email and mail, which likely contributed to the low response rate.^23^ In response, we shifted to community-based strategies including attending local events and distributing flyers. This increased engagement but was resource-intensive. We collaborated with an institutional data team to obtain and refine datasets of potential participants from the [University name] Eye Center and the [University name] Diabetes Center, who still required telephone contact to determine eligibility. Among all eligible individuals, there was a 27% translation from contact to study enrollment, suggesting that future iterations of the program will require optimized outreach strategies.

The SEEN Program was well received. Participants, the research team, and key partners all reported high satisfaction. Among the 35 enrolled participants, retention to completion of the year-long program was high (71.4%). This rate is comparable to other patient navigation programs for chronic diseases (60-77%), suggesting that the SEEN Program is acceptable to participants and feasible for real-world implementation.^24–26^ Participants cited positive relationships with patient navigators as a key factor in their satisfaction with the program. Indeed, the relationship-based framework of patient navigation is an important element of successful programs.^27^

Effective patient navigation is time-intensive; in our study, individuals with social needs required more time and touchpoints than those without (102 vs 81 minutes; 18.4 vs 16.5 touches). This reflects the time-intensive nature of addressing practical concerns such as transportation and food insecurity.^28^ The exact role of a patient navigator varies across programs. Our model was similar to the one described by Loskutova et al. for general diabetes management, with navigators serving as a combined case manager and resource liaison.^9^ Their patient navigators made an average of 6.1 calls per patient over 9 months.^9^ Our patient navigation was more demanding, requiring an average of 18 touches per patient over 12 months. This reflects our specification of quarterly check-ins, with some participants requiring multiple voicemails before making telephone contact, as well as the high clinical and social complexity of our participants. Ultimately, navigators reported that the process was efficient and practical for integration into their standard workflow.

Both quantitative and qualitative evidence confirmed that we engaged a population with complex medical and social needs. Participants were primarily Black and Latine individuals with significant comorbidities. Most (73.3%) of the screened participants had identified social needs, and all individuals with DR had social needs. This reiterates the disproportionate burden of adverse social determinants of health which contribute to disparities in DR outcomes.^5,29^ The high rate of emergency department visits and hospitalizations further emphasizes the complex health needs of this cohort. Food insecurity and transportation were the most commonly reported needs. This reflects the intersection of poverty, systemic disinvestment in marginalized communities, and barriers to accessing consistent healthcare.^30^ Food insecurity can exacerbate diabetes progression through inconsistent nutrition, and transportation barriers may prevent timely follow-up care.^31^ The prevalence of these factors among our high-risk population reflects structural inequities that impede effective disease management. These findings reinforce that DR is not an isolated diagnosis but rather a manifestation of broader systemic inequities. Social determinants of health, structural racism, and inadequate healthcare access shape DR outcomes.^2–6^ Integrated social and medical interventions for DR prevention and management are needed to address these disparities.^32^

Our preliminary findings suggest that the program benefited participants. Consistent with improved health outcomes in other interventions utilizing a patient navigator model, no participants in our study experienced DR progression during the study period.^9,33,34^ More participants who completed the study (56%) than those who did not (20%) had a follow-up eye exam. This represents a significant increase for a cohort previously disengaged from ophthalmic care. Interestingly, study completers also had a slightly higher rate of appointment cancelations and no-shows (76% vs. 70%). Study completers received more check-ins and healthcare navigation support, which may have prompted them to schedule more appointments. While immediate adherence to appointments was not guaranteed, the higher proportion attending follow-up exams suggests that engagement with the patient navigator may have ultimately facilitated access to ophthalmic care for those who remained in the program.

Mean visual acuity remained stable, which was likely influenced by high baseline visual acuity leaving little room for improvement. Mean HbA1c decreased among study completers by 1.3 (9.8 to 8.5) from baseline to 12-month follow-up. This clinically significant improvement in glycemic control is associated with reduced risk of DR progression and is comparable to the effect size of pharmaceutical agents for diabetes treatment.^35,36^ Participants corroborated this improvement in diabetes control, attributing it to care coordination resources and accountability provided by regular check-ins with patient navigators. Patient navigators were also able to effectively address participants’ social needs. Only 2 (6.5%) identified needs remained unresolved or unmet. Qualitative interviews also revealed overwhelmingly positive reactions to the quality of interpersonal interactions as well as gratitude for both the tangible resources and lifestyle changes that the program facilitated.

Limitations of this pilot study include the small sample size and lack of a control group, which preclude definitive conclusions regarding program efficacy. A key strength of this study was the high level of participation among Black and Latine individuals, given the high prevalence of diabetes and DR among these populations.^4^ We also leaned into study team and key partner expertise, which enhanced program effectiveness and cultural appropriateness. Participants received expert eye care by the study PI, who is a vitreoretinal surgeon, and from the [University name] Eye Center. Likewise, PA-NH served as a true collaborative partner, offering expert insights gained through longstanding experience navigating the local population.

### Conclusion

In summary, the SEEN Program pilot addressed social needs among individuals at high risk for DR through a feasible and acceptable intervention. Our evaluation demonstrated success in engaging individuals who previously delayed care. Systemic change is needed to reduce preventable blindness from DR. In parallel, patient-centered interventions can work within the existing environment to address disparities in DR care and outcomes. By combining risk stratification, clinical evaluation, education, and locally embedded patient navigation, the SEEN Program offers a successful model for improving eye care utilization and health outcomes among vulnerable populations at high risk for DR-related vision loss.

## Meeting presentations

This work has been presented in part at the Association of Research in Vision and Ophthalmology (ARVO) annual meeting in May 2024 and May 2025, and the American Academy of Ophthalmology (AAO) annual meeting in October 2024.

## Data Availability

No data was generated by this study. The following existing data sources were used: qualitative interviews and medical records. Interview data are available upon request to the corresponding author. Medical records are not available due to HIPAA compliance reasons.

## Acknowledgements

This publication was made possible by Grant Number 1 K23 EY030530-01 from the National Eye Institute. This work has been supported in part by an unrestricted/challenge award to Yale University Eye Center from the Research to Prevent Blindness (RPB), Inc. The sponsor or funding organization had no role in the design or conduct of this research.

